# Artificial intelligence for detecting bipolar disorder in electronic health records of patients with affective diagnoses: a diagnostic accuracy study

**DOI:** 10.64898/2026.05.07.26352679

**Authors:** Eugenio Ferro, Natalia Castaño-Villegas, Manuel F. Esteban Cárdenas, Ana María Gómez Puentes, Carlos Torres-Delgado, Laura Ortiz Calderon, Katherine Monsalve, José Zea

**Author notes:** **Corresponding author:**, Postal address: Clínica Montserrat, Calle 134 # 17-71, Bogotá 110121, Colombia.

## Abstract

**Background:** Bipolar disorder (BD) is frequently underdiagnosed, particularly in patients presenting with depressive disorders, leading to delays in appropriate treatment. Artificial intelligence (AI) applied to electronic health records (EHRs) may improve early detection by identifying clinically relevant symptom patterns.

**Objective:** To evaluate the diagnostic performance of a natural language processing (NLP)-based AI model for detecting BD-related features in EHRs of patients with affective diagnoses.

**Methods:** A retrospective diagnostic accuracy study was conducted using 500 EHRs from a psychiatric referral hospital in Bogotá, Colombia (2020–2024). The model extracted 18 predefined clinical domains from unstructured text and classified patients into four risk categories. Diagnostic performance was assessed in a validation subset of 100 records using independent psychiatric evaluation as the reference standard. Sensitivity, specificity, positive and negative predictive values, F1-score, and area under the receiver operating characteristic curve (AUC-ROC) were calculated.

**Results:** The model achieved high agreement in symptom extraction (mean 91.1%). Sensitivity was 96.4% (95% CI: 87.7%–99.0%) and specificity was 84.4% (95% CI: 71.2%–92.3%), with an F1-score of 0.92 and an AUC-ROC of 0.932 (95% CI: 0.881–0.975). A substantial proportion of patients with depressive diagnoses were identified as having confirmed BD or clinically relevant risk. The model analyzed complete EHRs 120 times faster than human reviewers.

**Conclusions:** NLP-based analysis of EHRs can achieve clinically meaningful performance in identifying BD-related patterns while substantially reducing review time. The model may be useful as a clinical decision support tool for earlier identification of bipolar disorder.

## 1. Introduction

Bipolar disorder (BD) is a complex, severe, chronic, and recurrent psychiatric condition, considered one of the most disabling mental disorders worldwide. Its global prevalence is estimated at approximately 2%, increasing to 4.4%–5% when conceptualized as part of the bipolar spectrum [1, 2]. BD is characterized by alternating hypomanic, manic, mixed, and depressive episodes, with a longitudinal predominance of depressive symptoms [2, 3]. The burden of BD is substantial, including increased mortality, persistent functional impairment even during remission, and a high lifetime disability load [2, 3]. Suicide risk is markedly elevated compared to the general population, with up to 20% of patients dying by suicide [2, 4]. In addition, depressive symptoms account for the majority of longitudinal morbidity, representing nearly three-quarters of symptomatic time in many patients [2, 3].

Despite its clinical severity and impact, BD remains significantly underdiagnosed in routine practice. Although symptom onset typically occurs in late adolescence or early adulthood, diagnosis is often delayed for several years, particularly when the initial presentation is depressive [2, 3, 5]. Contemporary evidence shows that diagnostic delay is especially prolonged in bipolar II disorder and that a substantial proportion of patients initially receive alternative diagnoses, most commonly within the depressive or anxiety spectrum [2, 3, 5].

Major depressive disorder (MDD) represents one of the primary sources of diagnostic masking of BD. Longitudinal studies have demonstrated clinically relevant rates of diagnostic conversion from MDD to BD, particularly among younger individuals and those with higher baseline psychiatric burden [6, 7]. Additional contributors to diagnostic delay include symptom overlap with other psychiatric conditions such as anxiety disorders, attention-deficit/hyperactivity disorder (ADHD), and borderline personality disorder [5, 8].

Delayed diagnosis is associated with significant clinical and socioeconomic consequences, including persistent functional impairment, poor social adjustment, increased frequency and severity of affective episodes, reduced treatment response, elevated suicide risk, and substantial economic burden [1–3]. These costs are largely driven by indirect factors such as loss of productivity, unemployment, disability, and repeated healthcare utilization [2, 3]. These challenges highlight the need for early identification of clinical patterns suggestive of BD, including affective lability, irritability, anxiety, and bipolar depressive features, using complementary tools beyond conventional diagnostic approaches.

In this context, artificial intelligence (AI), particularly machine learning and natural language processing (NLP), has emerged as a promising approach in mental health, enabling the analysis of large volumes of clinical data and the identification of complex patterns not readily detectable by human observers [9, 10]. Electronic health records (EHRs), which integrate both structured and unstructured clinical information, represent a valuable data source for such approaches.

Previous studies have reported accuracies ranging from 90% to 97% in the detection of depression and schizophrenia, and area under the curve (AUC) values up to 0.93 for suicide risk prediction using NLP applied to clinical notes [9]. However, the application of AI in mental health also presents important methodological and ethical challenges. Algorithms may reproduce implicit biases present in the data, potentially reinforcing disparities related to race, gender, or socioeconomic status [10, 11]. In addition, limited representation of Latin American populations restricts the external validity of many existing models [12].

To address these challenges, recent frameworks emphasize the integration of explainable AI (XAI), responsible AI principles, and human-in-the-loop validation to ensure transparency, fairness, and clinical applicability [10]. In Latin America, and particularly in Colombia, where access to mental health services is limited and specialized professionals are scarce, AI-based tools may have a transformative impact. Automated analysis of clinical narratives within routine EHRs could reduce diagnostic delay, improve diagnostic accuracy, and optimize the use of healthcare resources. However, despite growing interest, there is limited evidence on the clinical validation of NLP-based AI models for the detection of unrecognized BD in real-world Spanish-language EHRs. Therefore, the aim of this study was to validate an NLP-based artificial intelligence model for the identification of bipolar disorder criteria in routine electronic health records of patients with affective diagnoses.

## 2. Methods

### 2.1. Study design and population

A retrospective observational diagnostic accuracy study was conducted and reported in accordance with the STARD 2015 and TRIPOD+AI 2024 guidelines. Data were obtained from electronic health records (EHRs) of patients treated at Clínica Montserrat, University Hospital (ICSN), Bogotá, Colombia, between January 1, 2020, and December 31, 2024.

Records from patients aged ≥18 years with baseline diagnoses within the depressive or anxiety spectrum were included, with or without a prior diagnosis of bipolar disorder (BD). Diagnoses were defined using ICD-10 codes corresponding to affective and anxiety disorders, including F31.x (bipolar disorder), F32.x (depressive episode), F33.x (recurrent depressive disorder), and F412 (mixed anxiety-depressive disorder). Cases with a prior BD diagnosis were included for cohort characterization but were not used as the reference standard for diagnostic validation. Patients with active substance use disorders were excluded.

A total of 500 unique EHRs were selected by simple random sampling from a source population of 25,808 patients using a computer-generated random sequence. Sample size was estimated with OpenEpi version 3, assuming a BD prevalence of 25% in affective populations, a 5% margin of error, and a 95% confidence level, yielding a minimum required sample of 285 records. The final sample exceeded this threshold to improve the precision of the descriptive estimates.

For diagnostic accuracy analyses, a subsample of 100 EHRs was selected from the full cohort and evaluated against a clinical reference standard. The unit of analysis was the individual patient, and all available longitudinal information within each EHR was analyzed without segmentation by clinical episode. The selection process and analytical subsets are shown in Figure 1.

**Figure 1.**
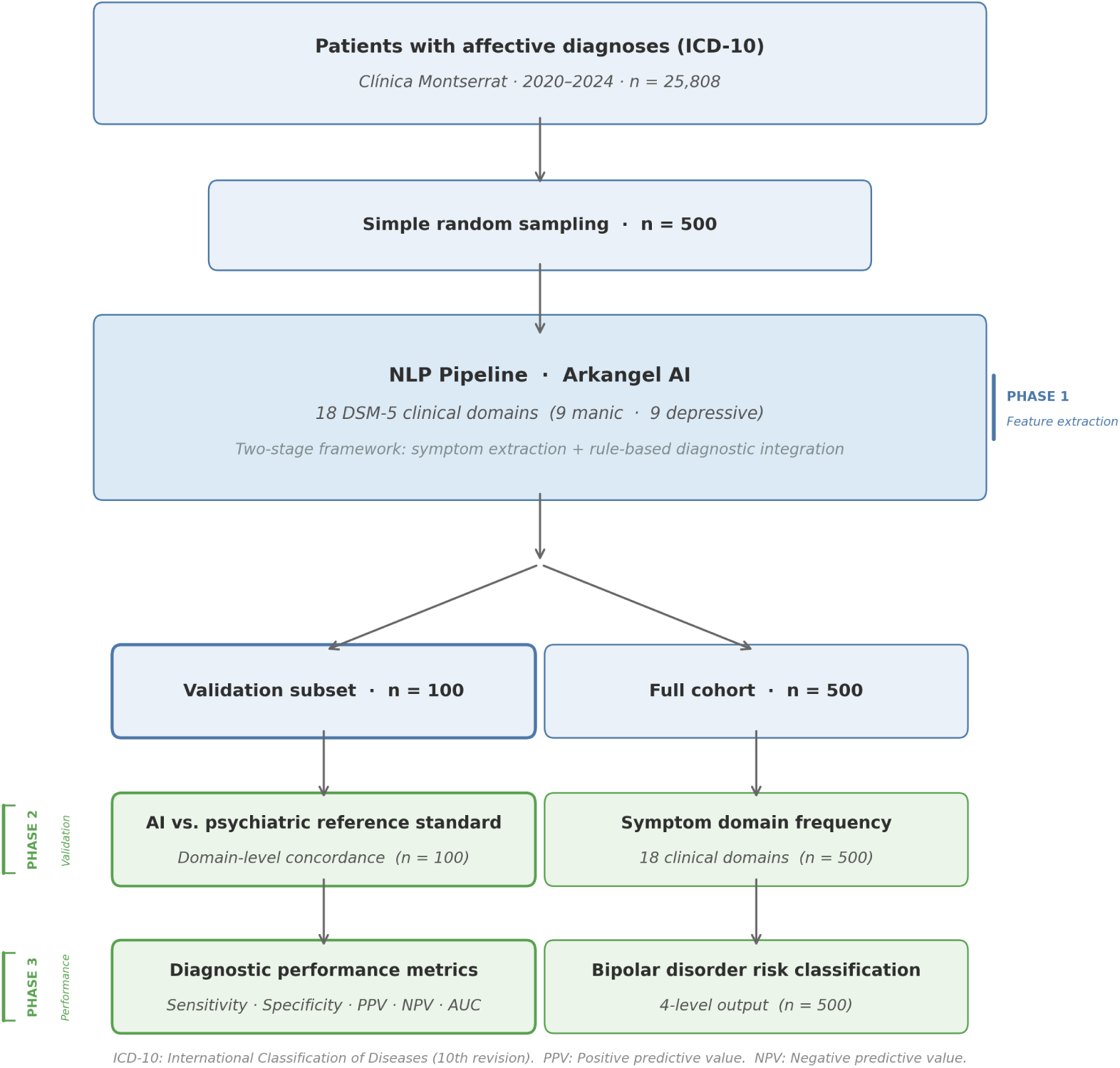
Participant selection flowchart according to STARD 2015.

### 2.2. Model application and clinical validation

The Arkangel AI model was designed as a two-stage system that separated clinical information extraction from diagnostic inference. In the first stage, 18 predefined clinical domains (nine manic and nine depressive) were extracted from unstructured clinical text. These domains were based on DSM-5 diagnostic criteria and the Mini International Neuropsychiatric Interview (MINI) [13].

Extraction was implemented using 18 independent prompts, each targeting a specific clinical domain and applying a guided chain-of-thought (CoT) reasoning strategy [14] (Supplementary Material 1). Each prompt produced a binary output (present/absent), enabling the construction of a structured patient-level dataset.

The model development included an iterative refinement phase based on an initial pilot using the first 30 EHRs. During this phase, the initial version of the 18 prompts was applied, and outputs were reviewed by psychiatrists to identify errors in symptom extraction, including omissions, false positives, and difficulties in interpreting implicit clinical narratives. Based on this evaluation, a first refinement was conducted, resulting in modifications to 15 of the 18 prompts, aimed at improving clinical sensitivity and semantic specificity. Three domains (decreased need for sleep, pressured speech, and flight of ideas) remained unchanged.

A second refinement phase introduced additional modifications to two domains (psychomotor retardation and suicidal ideation), along with adjustments to the clinical rules for identifying affective episodes, including the definition of temporal co-occurrence of symptoms. Following this process, both the prompts and the diagnostic integration rules were fixed, constituting the final version of the model. The complete set of final prompts is provided in Supplementary Material 1. All subsequent analyses, including the diagnostic accuracy evaluation, were performed with this final version, ensuring methodological consistency.

Model outputs were organized into a structured patient-level matrix, in which each clinical domain was represented as a binary variable (present/absent). Based on this representation, predefined rule-based criteria were applied to identify combinations of symptoms consistent with manic, hypomanic, and depressive episodes, allowing translation of unstructured clinical data into an interpretable and reproducible diagnostic framework.

Four diagnostic categories were defined: (1) confirmed BD, when DSM-5 criteria for bipolar disorder were met; (2) high probability of BD, defined as the presence of at least three manic symptoms within the same documented clinical episode; (3) some probability of BD, defined as the presence of three or more manic symptoms across the clinical history without evidence of temporal co-occurrence; and (4) no risk, in the absence of these criteria.

For diagnostic accuracy evaluation, the index test corresponded to the Arkangel AI model, and the reference standard consisted of independent clinical assessment by psychiatrists applying DSM-5 criteria and the MINI. Evaluators were trained prior to the study to standardize the application of diagnostic criteria. Each EHR was initially assessed by a primary reviewer; in cases of diagnostic uncertainty, a second independent evaluation was performed. Persistent discrepancies were resolved by a third independent reviewer, with the final diagnosis established by majority consensus. Reviewers were blinded to the model outputs.

Model validation was conducted in two stages. First, agreement in symptom extraction between the model and clinicians was evaluated at the domain level. Second, diagnostic performance was assessed by comparing model classifications with the clinical reference standard.

### 2.3. Statistical analysis

Sociodemographic and clinical variables were summarized using absolute and relative frequencies for categorical variables and measures of central tendency and dispersion for continuous variables, according to data distribution. Normality was assessed with the Shapiro–Wilk test. Statistical significance was defined as *α*= 0.05. Missing data were not imputed; denominators for each variable reflect the number of patients with available data. The frequency of clinical domains identified by the model was expressed as proportions of the total number of records. Agreement between the model and the clinical reference standard in symptom extraction was evaluated using Cohen’s kappa (*κ*), interpreted according to the Landis and Koch criteria [15]. Given the potential effect of high prevalence on *κ* values, Gwet’s AC_1_ coefficient was additionally calculated as a complementary measure [16].

For diagnostic performance analysis, a binary classification was adopted, defining a positive result as any model-generated alert (confirmed BD, high probability, or some probability) and a negative result as no risk, consistent with the model’s screening purpose. A 2×2 contingency table was constructed, and sensitivity, specificity, positive predictive value (PPV), negative predictive value (NPV), overall accuracy, and F1-score were calculated. Sensitivity was defined as the primary performance metric. In addition, category-specific one-vs-rest analyses were performed for each of the four diagnostic classes in the validation subset; these results are reported in Supplementary Material 2.

Confidence intervals (95%) for proportions were estimated using the Wilson method [17]. The area under the receiver operating characteristic curve (AUC-ROC) was estimated using bootstrap resampling with 2,000 iterations. All analyses were conducted using Python (version 3.9.6), utilizing the pandas, scikit-learn, SciPy, and statsmodels libraries.

### 2.4. Bias control

Measures were implemented to reduce potential sources of bias. Simple random sampling was used to reduce selection bias. Blinding of psychiatrist evaluators to model outputs reduced review and information bias during reference classification. The use of an independent clinical reference standard based on DSM-5 and MINI criteria reduced incorporation bias by separating model predictions from the diagnostic benchmark. Consensus review of discrepant cases was used to improve internal consistency of the reference standard. Nonetheless, because the study was conducted in a single specialized referral hospital, some degree of spectrum bias may persist and limit generalizability to other healthcare settings.

### 2.5. Ethical considerations

The study was conducted in accordance with the Declaration of Helsinki, CIOMS guidelines, and Resolution 8430 of 1993 of the Colombian Ministry of Health. It was classified as minimal-risk research. Given its retrospective design and the use of anonymized secondary data without direct patient intervention, informed consent was waived. The study protocol was approved by the Institutional Research Ethics Committee of Clínica Montserrat (ICSN) (approval number: Act No. 228, December 5, 2025).

## 3. Results

### 3.1. Cohort characteristics

A total of 500 patients with affective diagnoses treated between 2020 and 2024 were included. The median age was 31 years (IQR: 22–45), and 70.8% were female. Most patients were single (66.8%), and 62.8% had completed university-level education. Psychiatric family history was available in 91.4% of cases (n = 457), with depression being the most frequently reported condition (21.0%), followed by bipolar disorder (9.6%) and mania or hypomania (6.3%). The complete sociodemographic and clinical characteristics of the cohort are presented in Table 1.

**Table 1.** Baseline characteristics of the cohort.

| Variable | n (%) |
| --- | --- |
| **Age (years)** |  |
| Median [IQR] | 31 [21–44] |
| Minimum–Maximum | 18–90 |
| **Sex** |  |
| Female | 354 (70.8) |
| Male | 146 (29.2) |
| **Marital status** |  |
| Single † | 334 (66.8) |
| Married | 120 (24.0) |
| Cohabiting | 46 (9.2) |
| **Education** |  |
| University | 314 (62.8) |
| High school/Secondary education | 109 (21.8) |
| Postgraduate | 50 (10.0) |
| Technical/Technological training | 21 (4.2) |
| Primary education | 6 (1.2) |
| **Psychiatric family history ‡ (n = 457)** |  |
| Depression | 96 (21.0) |
| Bipolar disorder | 44 (9.6) |
| Mania/Hypomania | 29 (6.3) |

† Includes separated, divorced, and widowed individuals. ‡ Family psychiatric history data were unavailable for 43 patients in the clinical source; percentages were calculated over $n = 457$ . IQR: interquartile range. Age distribution was non-normal (Shapiro–Wilk $W = 0.887$ ; $p < 0.001$ ).

### 3.2. Symptom extraction by the model

The Arkangel AI system extracted all 18 predefined clinical domains across the 500 analyzed records. Among manic symptoms, irritability/hostility (84.0%), distractibility (77.2%), impulsivity (67.2%), and increased activity (60.6%) were the most frequent, whereas elevated mood (24.4%) and decreased need for sleep (16.4%) were the least frequent.

Across depressive symptoms, all domains were identified in more than 60% of cases. The most frequent were insomnia or hypersomnia (96.8%), cognitive symptoms (94.2%), anhedonia (92.6%), and depressed mood (91.8%). The complete frequency distribution by clinical domain is presented in Table 2.

**Table 2.** Frequency of clinical domains identified by the model in the full cohort (n = 500)

| Clinical domain | n | % |
| --- | --- | --- |
| **Manic/hypomanic symptoms** |  |  |
| Irritability/Hostility * | 420 | 84.0 |
| Distractibility/Labile attention | 386 | 77.2 |
| Risky activities/Impulsivity | 336 | 67.2 |
| Increased activity/Psychomotor agitation | 303 | 60.6 |
| Elevated/Expansive mood * | 122 | 24.4 |
| Pressured speech/Logorrhea | 87 | 17.4 |
| Decreased need for sleep | 82 | 16.4 |
| Flight of ideas/Racing thoughts | 77 | 15.4 |
| Grandiosity/Inflated self-esteem | 76 | 15.2 |
| **Depressive symptoms** |  |  |
| Insomnia or hypersomnia | 484 | 96.8 |
| Cognitive impairment (concentration/decision-making) | 471 | 94.2 |
| Anhedonia/Loss of interest or pleasure * | 463 | 92.6 |
| Depressed mood * | 459 | 91.8 |
| Feelings of worthlessness or excessive guilt | 437 | 87.4 |
| Thoughts of death/Suicidal ideation | 434 | 86.8 |
| Fatigue or loss of energy | 411 | 82.2 |
| Weight/appetite changes | 400 | 80.0 |
| Psychomotor agitation or retardation | 322 | 64.4 |

\* Core symptom required by the model’s diagnostic logic. Domains are ordered from highest to lowest frequency within each group. Percentages were calculated over the total sample (n = 500). A single patient could present multiple domains simultaneously.

### 3.3. Agreement in symptom extraction

The model demonstrated high agreement with clinical review across most domains. Among manic or hypomanic symptoms, observed agreement exceeded 84% in all domains, with the highest values for pressured speech (97.0%; *κ* = 0.891), grandiosity (97.0%; *κ* = 0.853), and irritability/hostility (96.0%; *κ* = 0.891). For depressive symptoms, agreement exceeded 90% in most domains, with the highest values for feelings of worthlessness or guilt (96.0%; *κ* = 0.779), thoughts of death or suicidal ideation (95.0%; *κ* = 0.787), and cognitive symptoms (95.0%; *κ* = 0.809). Lower *κ* values were observed in specific domains, particularly depressed mood (*κ* = 0.177) and insomnia or hypersomnia (*κ* = 0.468), in the context of high prevalence. Full agreement results by domain are presented in Table 3.

**Table 3.**
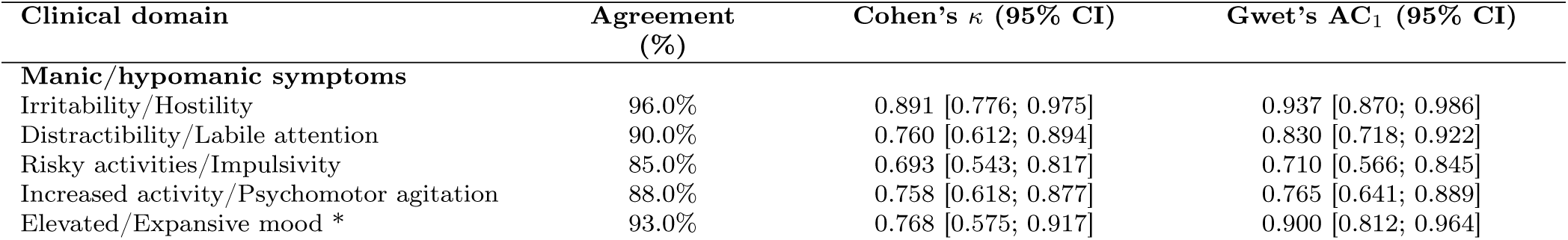
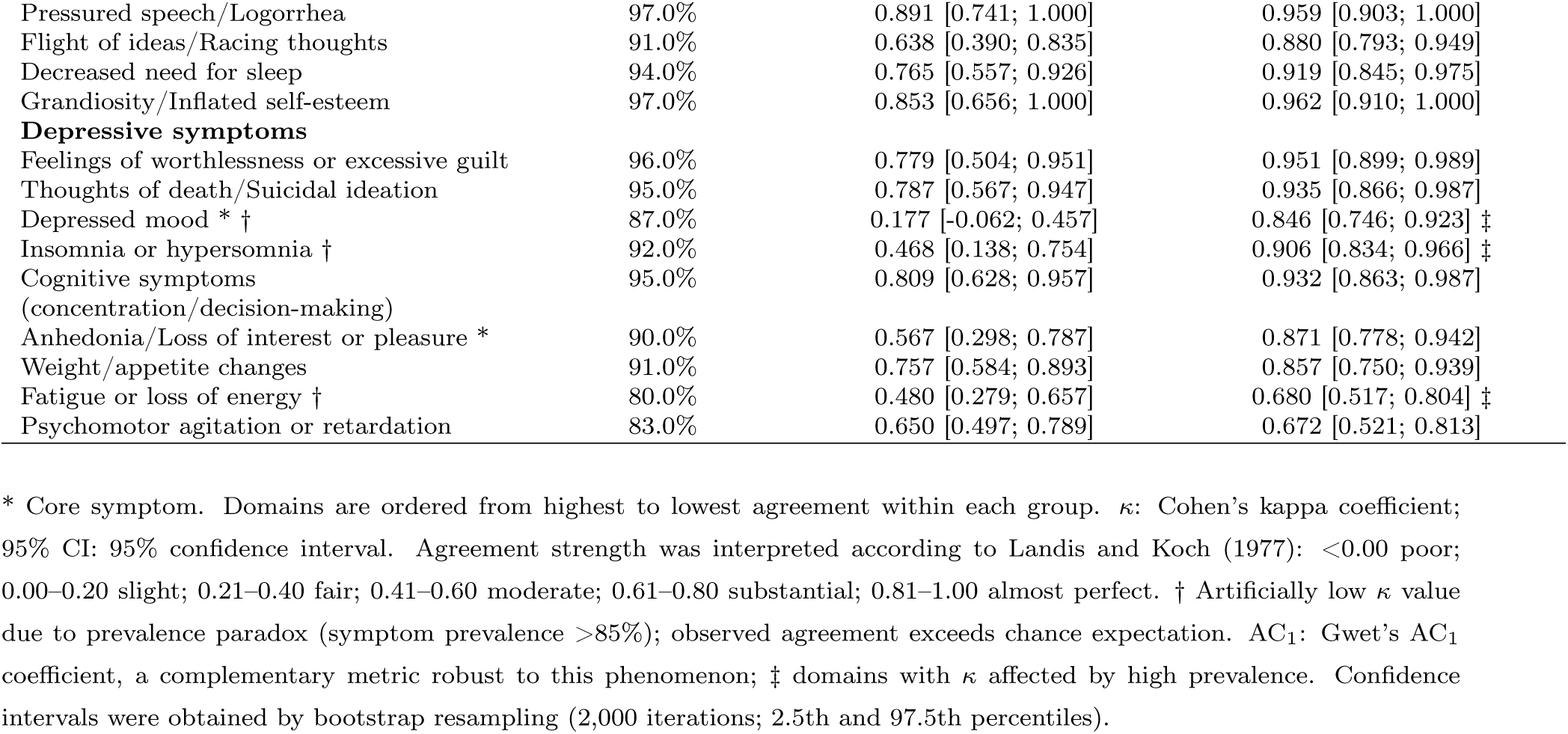
Agreement by clinical domain between the model and human review in the validation subset (n = 100)

### 3.4. Diagnostic performance

The model identified 53 true positives and 38 true negatives, with 7 false positives and 2 false negatives. Sensitivity was 96.4% (95% CI: 87.7%–99.0%), and specificity was 84.4% (95% CI: 71.2%–92.3%). The positive predictive value was 88.3%, and the negative predictive value was 95.0%. Overall accuracy was 91.0%, with an F1-score of 0.922 and an area under the receiver operating characteristic curve (AUC-ROC) of 0.932 (95% CI: 0.881–0.975). Complete results are presented in Table 4.

**Table 4.** Diagnostic performance of the model in the validation subset (n = 100)

| **A. Contingency table** |  |  |  |
| --- | --- | --- | --- |
|  | **Gold standard**<br>**positive †** | **Gold standard**<br>**negative ‡** | **Total** |
| Model positive § | 53 (TP) | 7 (FP) | 60 |
| Model negative § | 2 (FN) | 38 (TN) | 40 |
| **Total** | **55** | **45** | **100** |
| **B. Model operating characteristics** |  |  |  |
| **Metric** | **Value** | **95% CI** |  |
| Sensitivity | 96.4% | [87.7%; 99.0%] |  |
| Specificity | 84.4% | [71.2%; 92.3%] |  |
| PPV | 88.3% | [77.8%; 94.2%] |  |
| NPV | 95.0% | [83.5%; 98.6%] |  |
| Accuracy | 91.0% | [83.8%; 95.2%] |  |
| F1-score | 0.922 | — |  |
| AUC-ROC | 0.932 | [0.881; 0.975] |  |

TP = true positive; FP = false positive; FN = false negative; TN = true negative. PPV = positive predictive value; NPV = negative predictive value; AUC-ROC = area under the receiver operating characteristic curve. 95% CIs were calculated using the Wilson method (proportions) and bootstrap resampling (2,000 iterations; AUC-ROC). † Positive gold standard: psychiatric verdict of confirmed BD, high probability, or some probability. ‡ Negative gold standard: no risk. §Positive model: classification of confirmed BD, high probability, or some probability; negative model: no risk. F1 = harmonic mean of precision and sensitivity, where 1.0 is the maximum value.

### 3.5. Bipolar disorder risk classification

A similar overall distribution was observed between the classifications generated by the Arkangel AI model and those assigned by the clinical reference standard. In both, the most frequent category was “no risk” (model: n = 40; clinical standard: n = 45), followed by confirmed bipolar disorder (model: n = 25; clinical standard: n = 26). In the intermediate categories, the model classified more cases as “some probability” (n = 25 vs. 17), whereas the clinical reference standard identified more cases as “high probability” (n = 12 vs. 10). The comparative distribution of classifications is shown in Figure 2.

**Figure 2.**
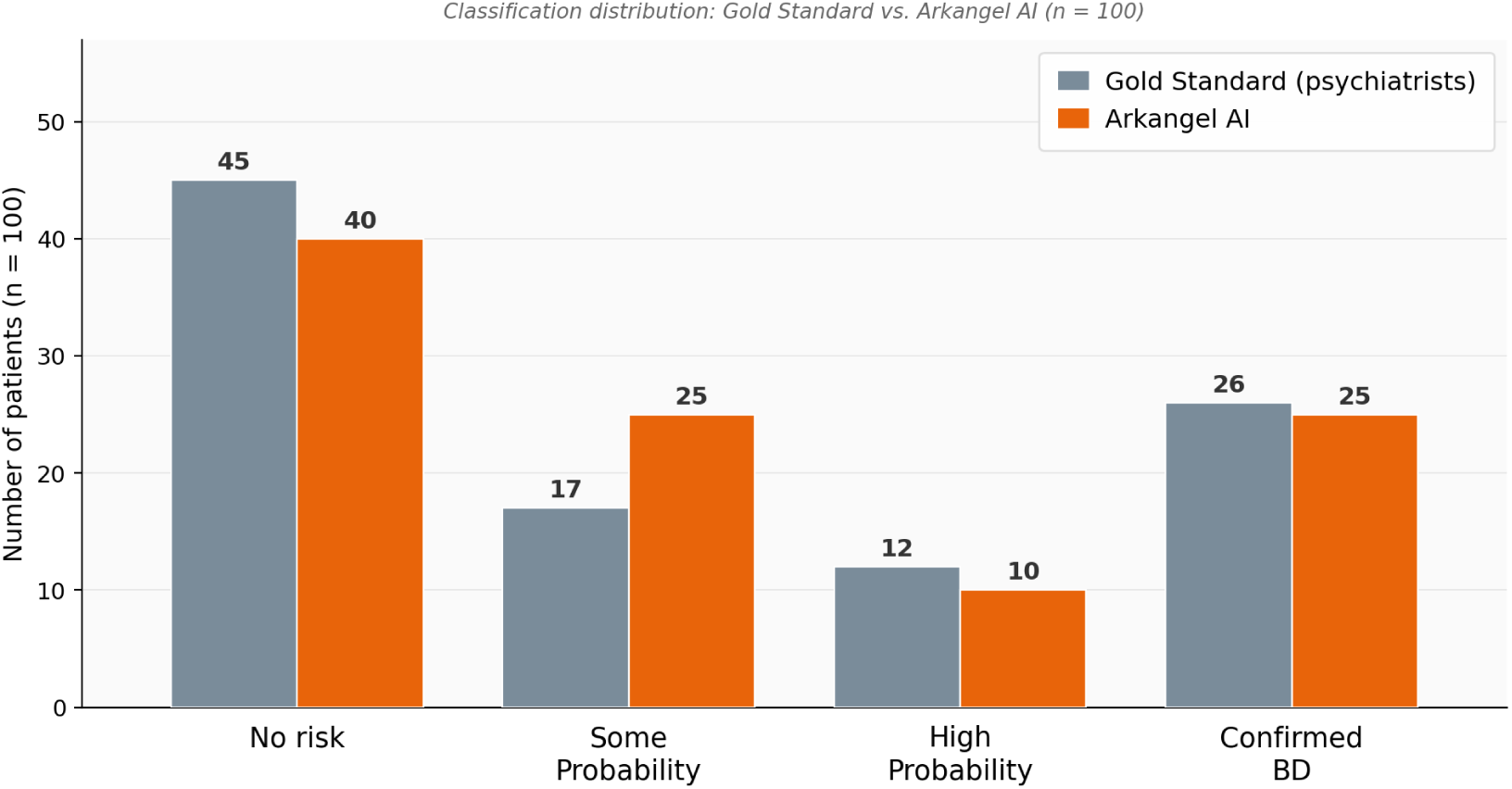
Distribution of bipolar disorder risk classification comparing the Arkangel AI model and the reference clinical standard in the validation subset (n = 100).

In the full cohort (n = 500), the most frequent category was “no risk” (n = 247; 49.4%), followed by confirmed bipolar disorder (n = 112; 22.4%). Intermediate risk categories corresponded to high probability (n = 90; 18.0%) and some probability (n = 51; 10.2%).

The distribution of classifications between the model and the clinical standard at the individual category level is presented in Figure 3. Category-specific one-vs-rest analyses are provided in Supplementary Material 2 and show that performance was strongest in the extreme categories (confirmed BD and no risk), whereas the greatest overlap occurred between the two intermediate risk categories.

**Figure 3.**
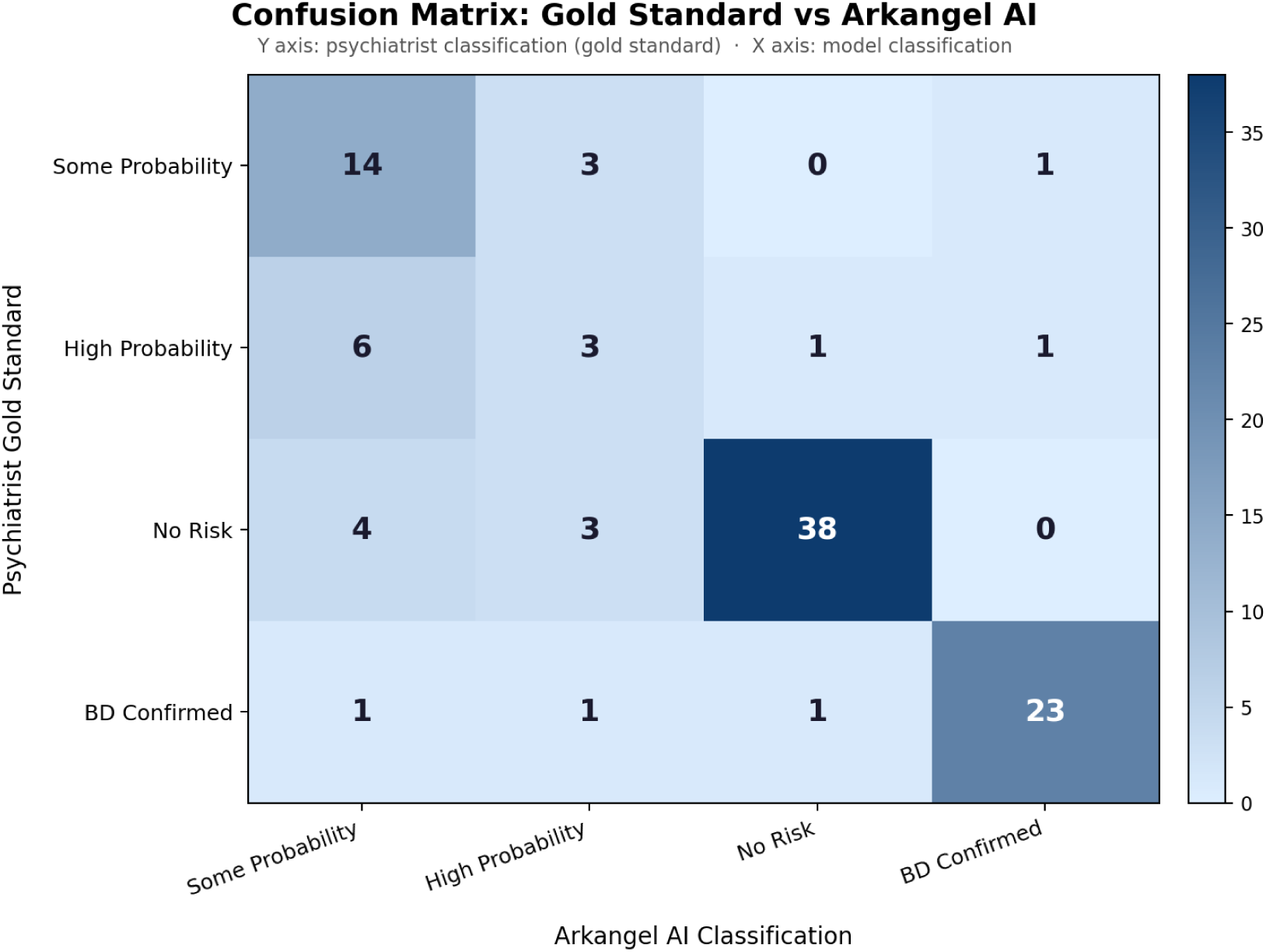
Multiclass confusion matrix comparing classifications generated by the Arkangel AI model with the reference clinical standard in the validation subset (n = 100).

### 3.6. Processing time analysis

In the validation subset (n = 100), human reviewers evaluated a total of 9,896 pages over 41.3 cumulative hours, with an average review time of 24.8 minutes per case. The Arkangel AI system processed the same set of clinical records in 20 minutes and 32 seconds, equivalent to approximately 0.2 minutes per case. This corresponds to a 99.2% reduction in total analysis time, an approximately 120-fold difference in time per case between human review and the AI system. Figure 4 illustrates the relationship between record length and human review time, as well as the comparison of time per case between both approaches.

**Figure 4.**
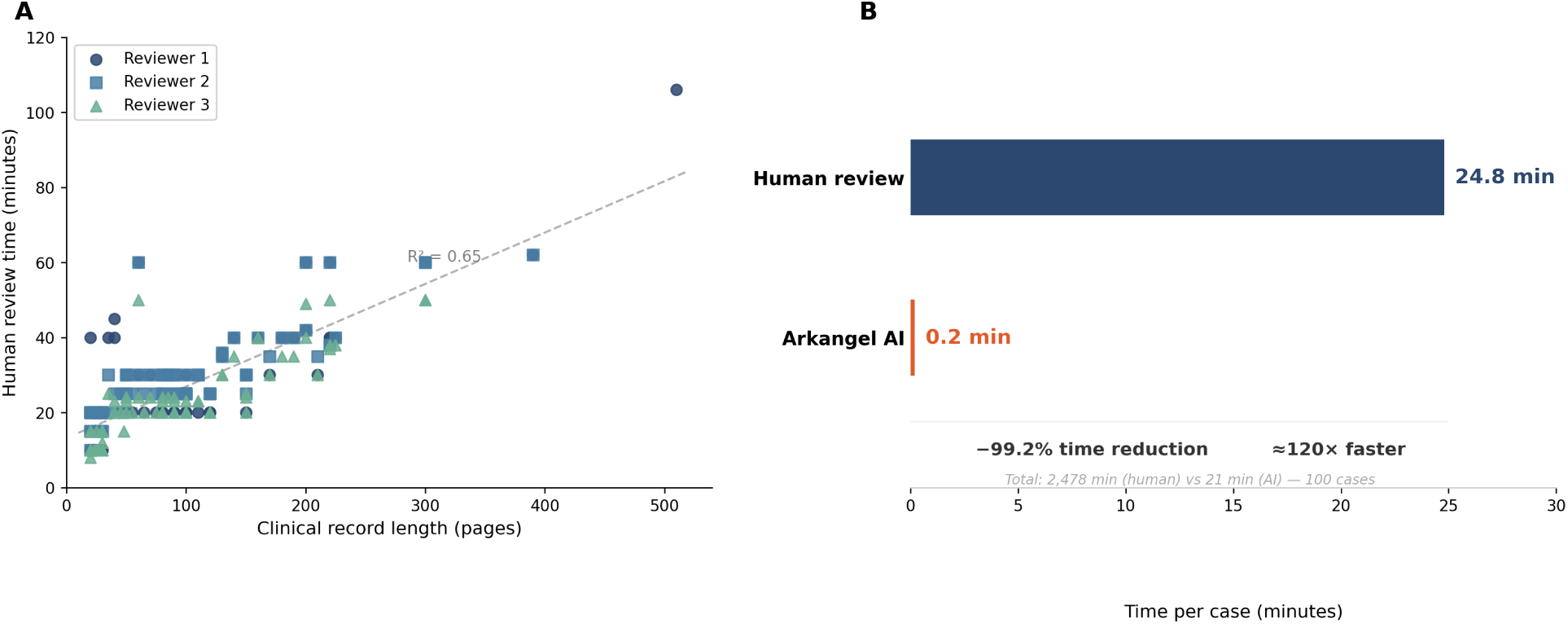
Comparative analysis of review time in the validation subset (n = 100). Panel A shows the relationship between record length (pages) and human review time per case; each point represents one reviewer. Panel B compares time per case between human review and the Arkangel AI model.

## 4. Discussion

This study evaluated a natural language processing (NLP)-based artificial intelligence model for identifying BD-related symptom patterns in Spanish-language electronic health records from a psychiatric referral hospital in Bogotá, Colombia. The model achieved high agreement in symptom extraction (mean 91.1%) and strong binary screening performance in the validation subset, with sensitivity of 96.4%, specificity of 84.4%, an F1-score of 0.92, and an AUC-ROC of 0.932. Category-specific analyses further showed the strongest performance in the extreme classes, with F1-scores of 0.902 for confirmed BD and 0.894 for no risk, whereas discrimination was weaker in the intermediate categories, particularly high probability (F1-score 0.273) (Supplementary Material 2). In the full cohort, 22.4% of records were classified as confirmed BD, 18.0% as high probability, and 10.2% as some probability, while review time was reduced by 99.2% compared with psychiatrist assessment. Taken together, these findings support the potential value of AI-assisted screening in settings where bipolarity is frequently underrecognized and diagnostic delay remains substantial [18, 19].

These results are broadly consistent with the current literature, but comparisons should be interpreted in light of differences in study design and clinical task. Pan et al. reported pooled sensitivities of 0.84–0.88, specificities of 0.82–0.89, and AUC values of 0.89–0.94 across machine learning studies of BD diagnosis [20]. Colombo et al. described overall classification accuracies around 0.77 in BD prediction models using biological, clinical, and neuropsychological markers [21]. The AUC-ROC observed here (0.932) exceeds that reported by Hansen et al. (0.62) for predicting future diagnostic progression to BD [22], but the tasks are not equivalent: the present model performs retrospective identification of symptom patterns already documented in narrative EHRs, whereas Hansen et al. addressed prospective prediction of later diagnostic conversion. Additional NLP studies in psychiatry have reported F1-scores above 0.80 and diagnostic concordance levels comparable to those of early-career psychiatrists [23–25]. Particularly relevant is the work of De La Hoz et al., which demonstrated the feasibility of extracting psychiatric phenotypes from Spanish-language EHRs in Colombian hospitals [26]. In that context, the present study adds evidence from a clinically supervised, Spanish-language, real-world screening framework validated against independent expert psychiatric review in a Latin American psychiatric setting. This is also consistent with prior EHR phenotyping studies reporting positive predictive values near 0.85 compared with expert chart review [27].

A central finding is that the model’s greatest strength lies in symptom extraction rather than in categorical diagnostic integration. This interpretation is supported by the very high domain-level agreement for several clinically salient variables, including pressured speech (97.0%; *κ* = 0.891), irritability/hostility (96.0%; *κ* = 0.891), weight/appetite changes (91.0%; *κ* = 0.757), and suicidal ideation or thoughts of death (95.0%; *κ* = 0.787). At the same time, the concentration of misclassification in the intermediate categories, together with lower agreement for semantically broad or highly prevalent domains such as depressed mood and insomnia or hypersomnia, indicates that the main difficulty lies in determining how symptoms cluster, co-occur over time, and acquire diagnostic meaning. This distinction is clinically relevant because psychiatric diagnosis depends not only on the presence of symptoms but also on their temporal organization, severity, and contextual interpretation, as reflected in structured diagnostic frameworks for BD and in the differential diagnostic literature [8, 13].

During iterative refinement, including the pilot phase and early validation rounds, psychiatrists reviewing model outputs also performed a qualitative assessment of errors and borderline cases. This process added information that was not captured by summary performance metrics alone. Early versions of the model had difficulty identifying suicidal ideation when it was only indirectly documented, and they occasionally inferred additional symptoms once a diagnostic threshold had been reached, suggesting overgeneralization from partial textual cues. The model also showed limitations when interpreting culturally nuanced expressions or weighting symptom intensity and temporal relevance across longitudinal records. Although iterative refinement improved detection of non-explicit symptoms and reduced false-positive symptom attribution, some degree of overestimation persisted when isolated behaviors were interpreted as recurrent clinical features.

These observations underscore a broader limitation of NLP systems in psychiatry: the challenge is not only extracting explicit symptom mentions, but also interpreting their clinical significance within a longitudinal and contextual framework, marking a critical boundary between pattern recognition and interpretative clinical reasoning [12, 23, 28, 29]. Psychiatric narratives often depend on implicit, culturally situated, and context-dependent language that requires interpretative reasoning rather than pattern recognition alone [12, 23]. For that reason, the most appropriate role for systems such as this one is likely within a human-in-the-loop clinical decision support model, in which computational efficiency in reviewing large volumes of narrative text complements, rather than replace, psychiatric judgment [28, 29].

The clinical implications of these findings are particularly relevant given the persistent delay in BD diagnosis. Population-based and clinical studies have shown that diagnostic delay is common, often prolonged, and especially marked in patients whose initial presentation is depressive [18, 19]. In that context, a tool with high sensitivity and substantially reduced review time may be useful as a first-line screening aid for services that must triage large volumes of unstructured EHR data. Its most plausible use is not autonomous diagnosis, but prioritization of patients whose records contain symptom constellations compatible with bipolarity and who may benefit from structured interview, longitudinal review, or specialist reassessment. This may be especially valuable in resource-constrained settings, where specialist time is limited and documentation burden is high. It may also have relevant implications for health system efficiency by optimizing allocation of specialized resources and reducing time to appropriate diagnostic evaluation. At the same time, the weaker performance observed in intermediate-risk categories supports cautious implementation and reinforces the need for clinician oversight in final diagnostic interpretation.

### 4.1. Strengths and limitations

This study has several strengths. It was conducted on real-world Spanish-language EHRs from a psychiatric hospital in Latin America, a context that remains underrepresented in the literature [12, 26]. Internal validity was strengthened by design features intended to reduce bias, including simple random sampling to mitigate selection bias, blinded psychiatrist review to reduce review and information bias, an independent DSM-5- and MINI-based reference standard to reduce incorporation bias, and consensus resolution of discrepant cases to improve the consistency of the final clinical classification. The model was also evaluated against expert psychiatric review rather than administrative coding alone, increasing the clinical relevance of the benchmark [27].

These strengths should be interpreted alongside several limitations. The retrospective design and single-center sample limit generalizability, particularly to non-specialized settings, and may introduce spectrum effects related to case complexity in a referral hospital. Reliance on routinely documented EHR narratives also creates a risk of information bias when symptoms are incompletely recorded, ambiguously phrased, or unevenly described across clinicians. The reference standard was based on independent evaluations by three psychiatrists, but interrater reliability was not formally quantified, so some observed disagreement may reflect variability in expert interpretation rather than model error alone. In addition, the model was developed and validated exclusively using Spanish-language free-text EHRs from a single Colombian institution and has not been tested in other languages, regions, or health information systems, limiting external applicability [23, 26]. Model performance may also be influenced by institution-specific documentation patterns, which could limit reproducibility in other clinical contexts. The absence of prospective follow-up precludes evaluation of whether patients classified as high probability or some probability would subsequently meet full diagnostic criteria for BD. Finally, the lower agreement observed in semantically broader domains and the concentration of disagreement in intermediate categories indicate that part of the remaining error stems from the inherent subjectivity of psychiatric interpretation rather than from symptom detection alone. These limitations support future multicenter studies with external validation, formal interrater reliability assessment, and prospective clinical follow-up.

## 5. Conclusion

This study provides evidence that NLP-based analysis of Spanish-language EHRs can achieve clinically meaningful performance in identifying BD-related symptom patterns while substantially reducing review time. The findings support the role of artificial intelligence as a complementary clinical decision support tool for prioritizing patients who may require more detailed bipolar disorder assessment, particularly in resource-constrained settings. Future research should focus on external validation, prospective implementation, and evaluation of clinical impact in real-world practice.

## Data Availability

De-identified data (including structured datasets and model outputs) are available upon reasonable request to the corresponding author, in accordance with institutional and ethical regulations. The analysis code is available from the corresponding author upon reasonable request.

## Acknowledgments

The authors would like to thank Clínica Montserrat for providing access to anonymized electronic medical records and for its support in conducting this study. The authors also thank the clinical team for their contribution to data collection and expert review.

## Author Contributions (CRediT)

N.C.V.: Conceptualization, Investigation, Data curation, Methodology, Validation, Writing – review & editing.

K.M.B.: Methodology, Formal analysis, Writing – original draft.

L.O.C.: Software, Data curation, Methodology, Validation, Writing – review & editing. A.M.G.P.: Resources, Writing – review & editing.

M.F.E.C.: Methodology, Resources, Writing – review & editing.

C.T.D.: Investigation, Writing – review & editing.

J.Z.: Resources, Supervision, Validation.

E.F.: Project administration, Methodology, Supervision, Resources, Visualization, Writing – review & editing.

All authors approved the final version of the manuscript.

## Data Sharing Statement

De-identified data (including structured datasets and model outputs) are available from the corresponding author upon reasonable request, in accordance with institutional and ethical regulations. The analysis code is available from the corresponding author upon reasonable request.

## Declaration of Interests

Some authors are employees of Arkangel AI, the developer of the artificial intelligence model evaluated in this study, and/or Clínica Montserrat, the institution where the data were collected. These affiliations are disclosed in the author information. The authors declare that the study was conducted independently and that the employers had no role in the study design, data analysis, interpretation of the results, or decision to submit the manuscript for publication.

## Role of the Funding Source

This research did not receive any specific grant from funding agencies in the public, commercial, or not-for-profit sectors. The artificial intelligence model evaluated in this study was developed by Arkangel AI within the context of an institutional collaboration with Clínica Montserrat.

## Declaration of Generative AI and AI-Assisted Technologies in the Manuscript Preparation Process

During the preparation of this manuscript, the authors used artificial intelligence tools (DeepL and GPT-4o, OpenAI) to assist with language translation and improve clarity of the text. All outputs were reviewed and edited by the authors, who take full responsibility for the content of the manuscript.

## Supplementary Material 1

English translation of the prompts used by the Arkangel AI extraction system (final implemented version)

This supplementary file presents an English translation of the final prompts used to extract the 18 predefined clinical domains from unstructured electronic health records. Because the model was implemented on Spanish-language records, the quoted example expressions are intentionally preserved in Spanish whenever they reflect the operational wording used in the original prompt set.

**Table S1.1.**
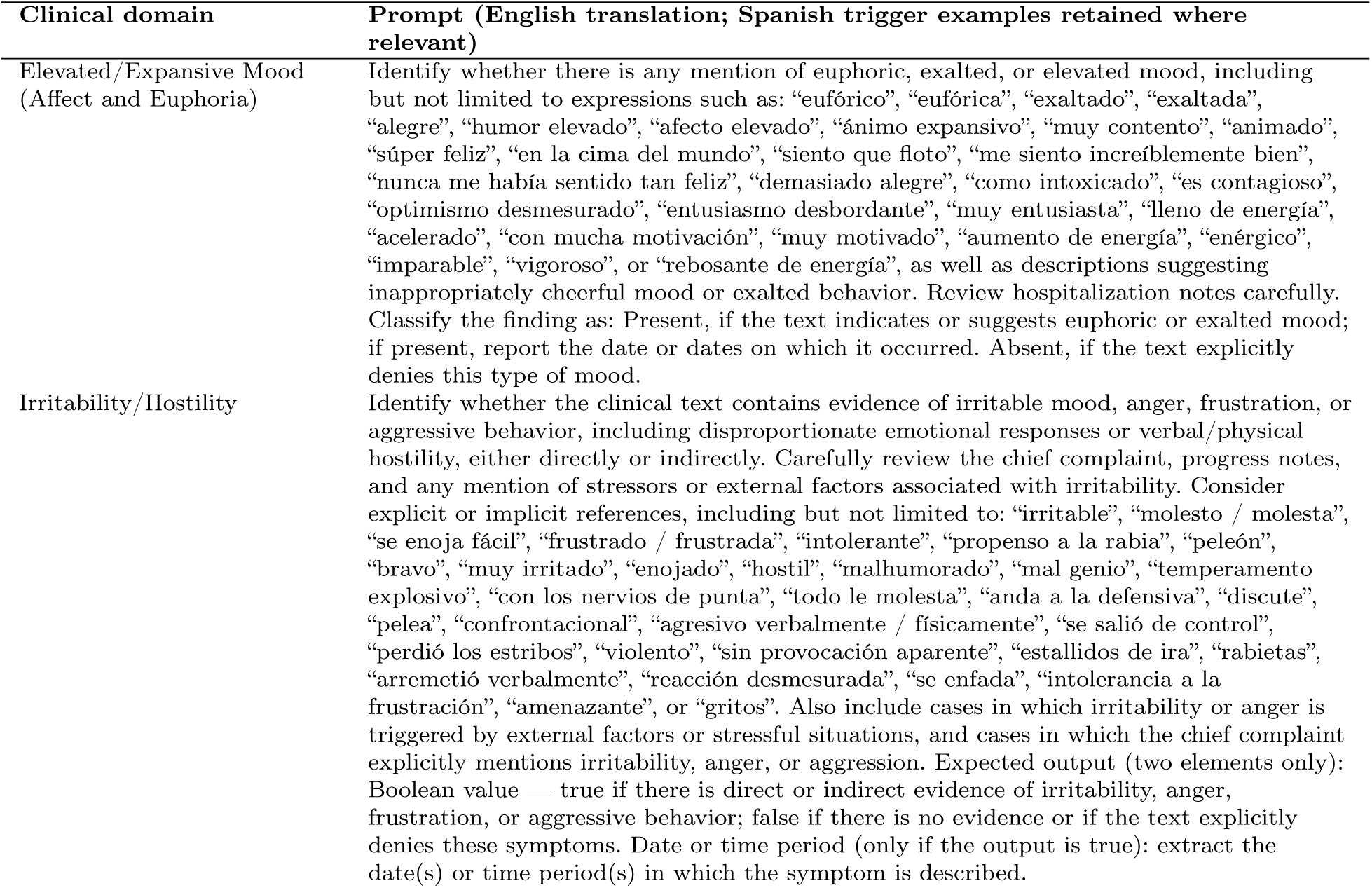
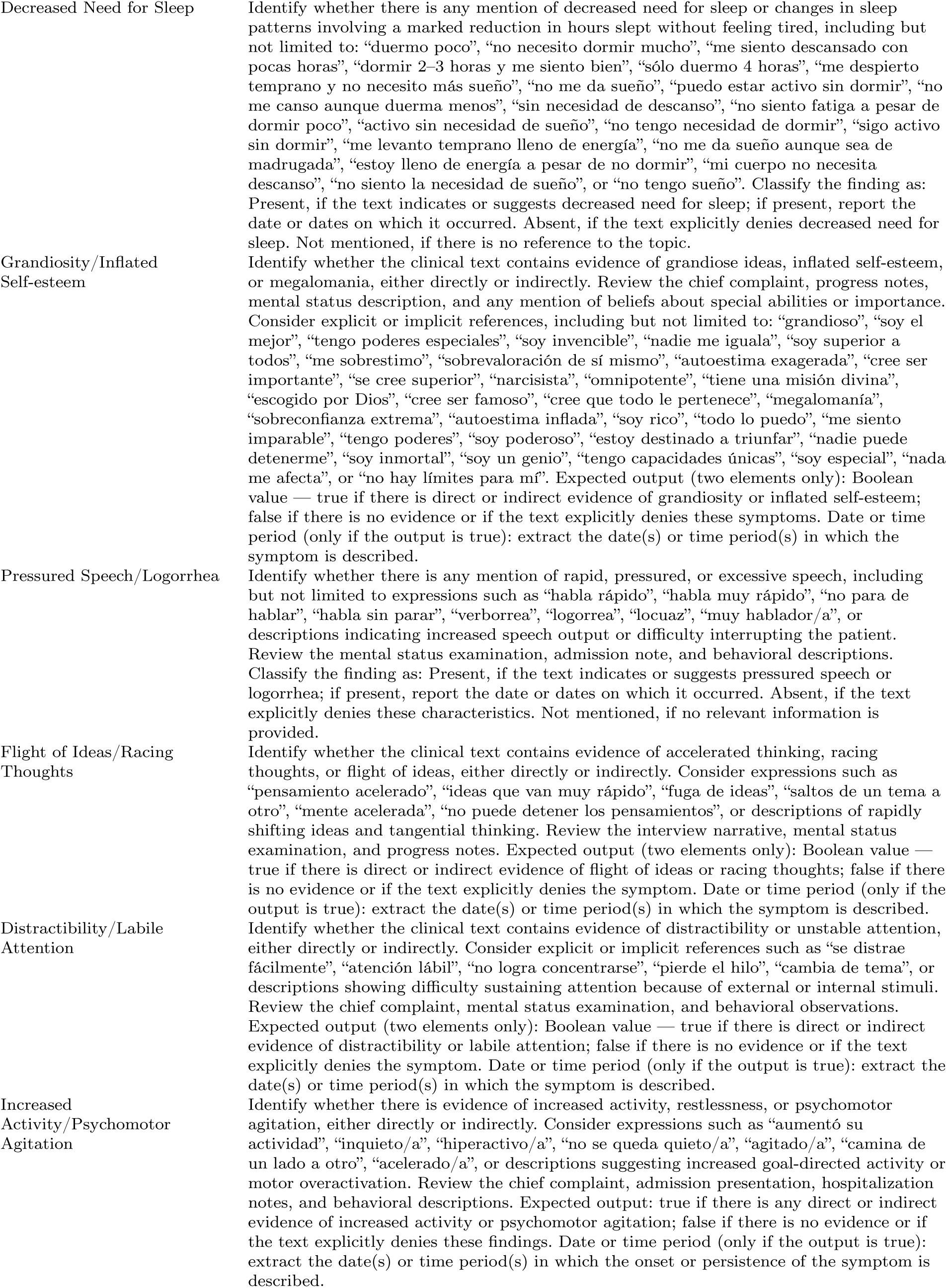
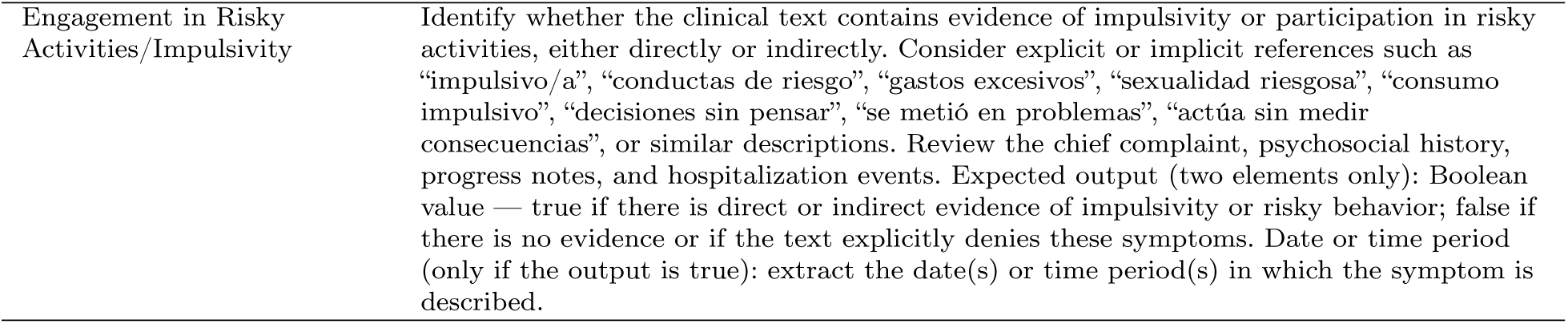
English translation of the final extraction prompts for manic and hypomanic symptoms (*n* = 9).

**Table S1.2.**
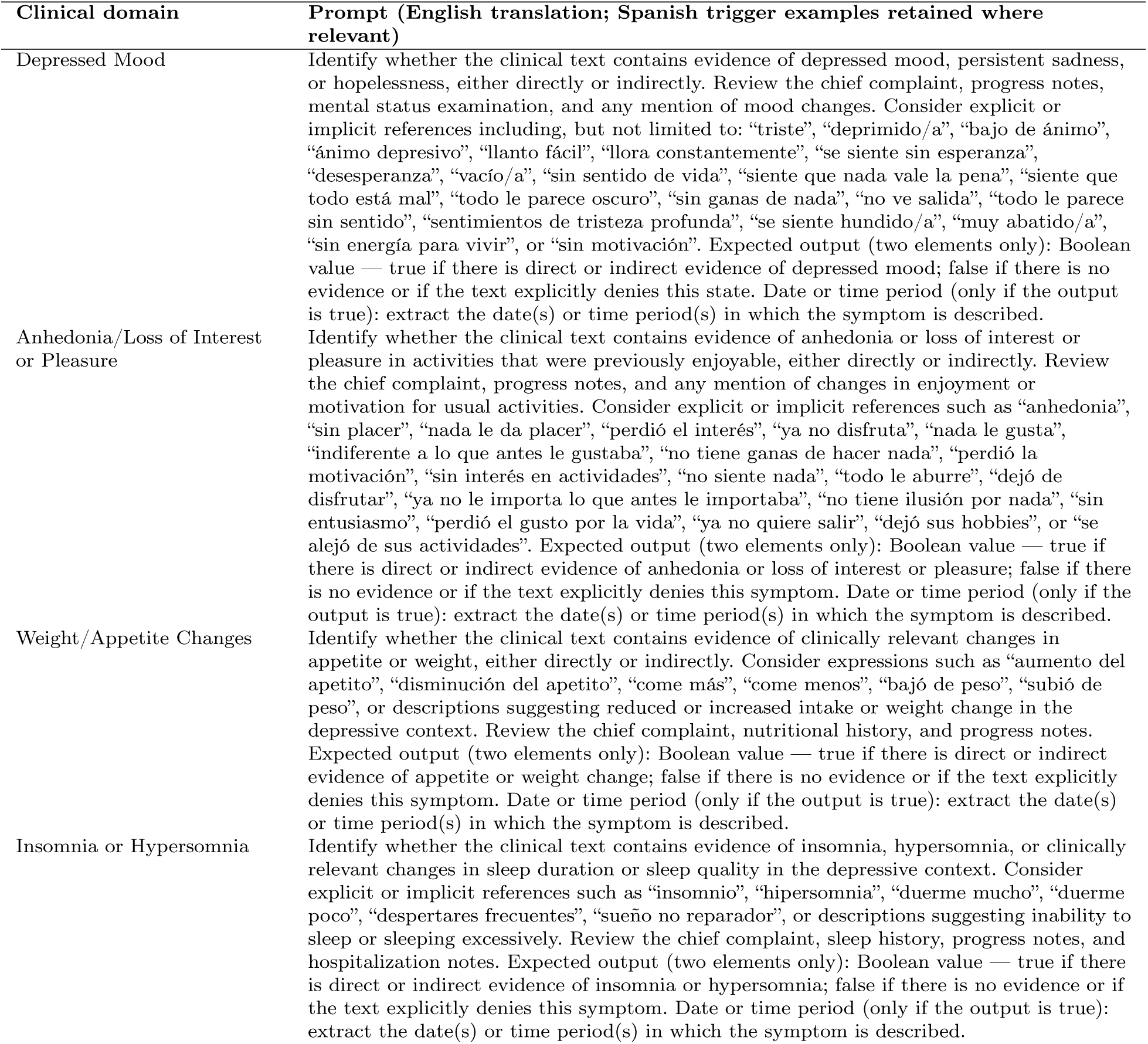
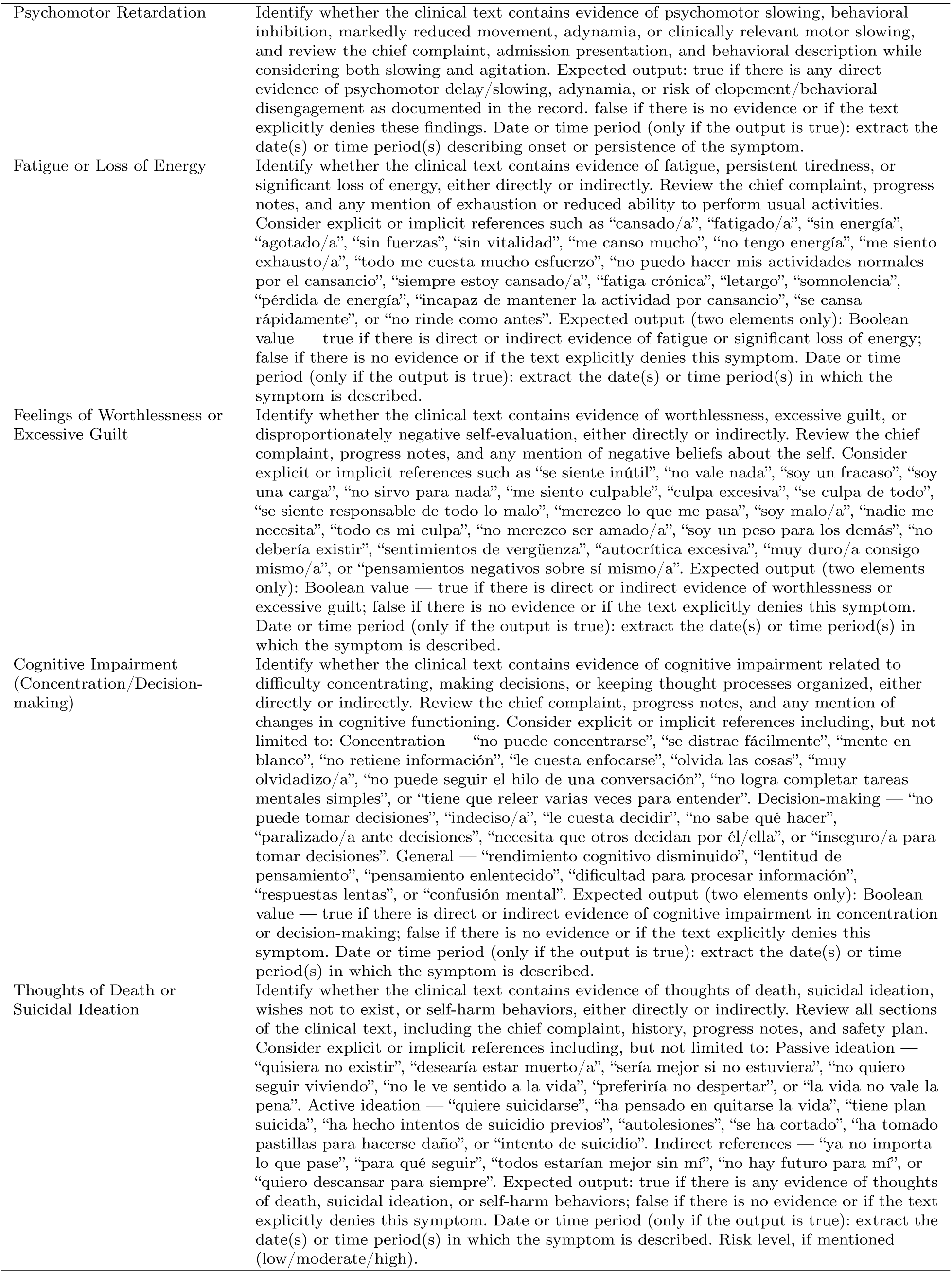
English translation of the final extraction prompts for depressive symptoms (*n* = 9).

## Supplementary Material 2

Category-specific analyses of bipolar disorder risk classification in the validation subset

All analyses in this supplementary file correspond to the validation subset (*n* = 100). Category-specific metrics were calculated using one-vs-rest (OvR) analyses, treating each category as positive and the remaining three categories as negative. Confidence intervals (95%) for proportions were calculated using the Wilson method. TP = true positive; FP = false positive; FN = false negative; TN = true negative; PPV = positive predictive value; NPV = negative predictive value.

**Table S2.1.** Full 4×4 confusion matrix for bipolar disorder risk classification in the validation subset (*n* = 100).

| Psychiatrist gold standard | Confirmed BD | High probability | Some probability | No risk | Total |
| --- | --- | --- | --- | --- | --- |
| Confirmed BD | 23 | 1 | 1 | 1 | 26 |
| High probability | 1 | 3 | 7 | 1 | 12 |
| Some probability | 1 | 3 | 13 | 0 | 17 |
| No risk | 0 | 3 | 4 | 38 | 45 |
| Total | 25 | 10 | 25 | 40 | 100 |

The pattern of discordance shows that most classification errors were concentrated between the two intermediate categories. In particular, 7 of the 12 cases classified by psychiatrists as high probability were assigned by the model to some probability, whereas the extreme categories (confirmed BD and no risk) showed few cross-category errors.

| Model vs psychiatrist | Gold + (Confirmed BD) | Gold - (rest) | Total |
| --- | --- | --- | --- |
| Model + (Confirmed BD) | TP = 23 | FP = 2 | 25 |
| Model - (rest) | FN = 3 | TN = 72 | 75 |
| Total | 26 | 74 | 100 |

**Table S2.2.** One-vs-rest analysis for Confirmed BD. Metric Value 95% CI.

| Metric | Value | 95% CI |
| --- | --- | --- |
| Sensitivity | 88.5% | [71.0%; 96.0%] |
| Specificity | 97.3% | [90.7%; 99.3%] |
| PPV (Precision) | 92.0% | [75.0%; 97.8%] |
| NPV | 96.0% | [88.9%; 98.6%] |
| F1-score | 0.902 | — |

| Model vs psychiatrist | Gold + (High probability) | Gold - (rest) | Total |
| --- | --- | --- | --- |
| Model + (High probability) | TP = 3 | FP = 7 | 10 |
| Model - (rest) | FN = 9 | TN = 81 | 90 |
| Total | 12 | 88 | 100 |

**Table S2.3.**
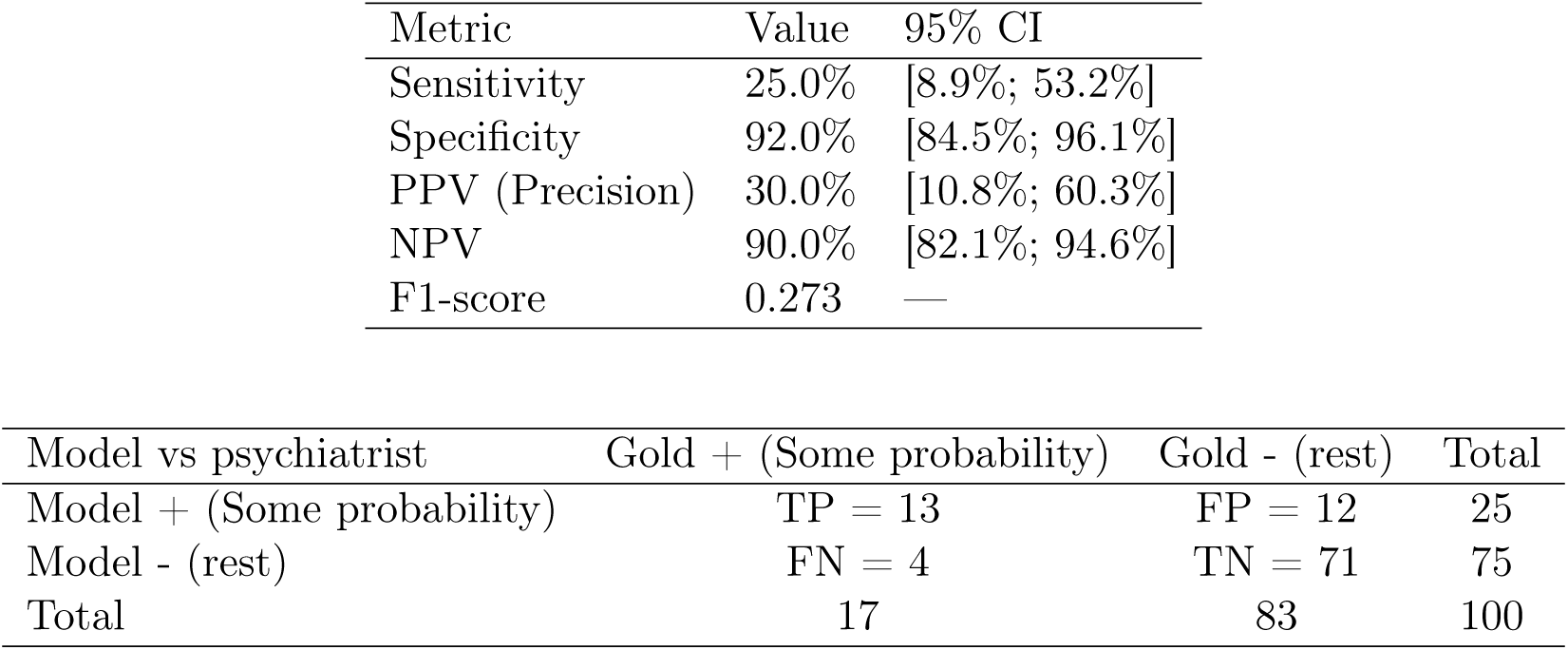
One-vs-rest analysis for High probability.

| Metric | Value | 95% CI |
| --- | --- | --- |
| Sensitivity | 25.0% | [8.9%; 53.2%] |
| Specificity | 92.0% | [84.5%; 96.1%] |
| PPV (Precision) | 30.0% | [10.8%; 60.3%] |
| NPV | 90.0% | [82.1%; 94.6%] |
| F1-score | 0.273 | — |

| Model vs psychiatrist | Gold + (Some probability) | Gold - (rest) | Total |
| --- | --- | --- | --- |
| Model + (Some probability) | TP = 13 | FP = 12 | 25 |
| Model - (rest) | FN = 4 | TN = 71 | 75 |
| Total | 17 | 83 | 100 |

**Table S2.4.** One-vs-rest analysis for Some probability.

| Metric | Value | 95% CI |
| --- | --- | --- |
| Sensitivity | 76.5% | [52.7%; 90.4%] |
| Specificity | 85.5% | [76.4%; 91.5%] |
| PPV (Precision) | 52.0% | [33.5%; 70.0%] |
| NPV | 94.7% | [87.1%; 97.9%] |
| F1-score | 0.619 | — |

| Model vs psychiatrist | Gold + (No risk) | Gold - (rest) | Total |
| --- | --- | --- | --- |
| Model + (No risk) | TP = 38 | FP = 2 | 40 |
| Model - (rest) | FN = 7 | TN = 53 | 60 |
| Total | 45 | 55 | 100 |

**Table S2.5.** One-vs-rest analysis for No risk.

| Metric | Value | 95% CI |
| --- | --- | --- |
| Sensitivity | 84.4% | [71.2%; 92.3%] |
| Specificity | 96.4% | [87.7%; 99.0%] |
| PPV (Precision) | 95.0% | [83.5%; 98.6%] |
| NPV | 88.3% | [77.8%; 94.2%] |
| F1-score | 0.894 | — |

**Table S2.6.** Summary of category-specific operating characteristics from one-vs-rest analyses.

| Category | *n* psychiatrist | *n* model | Sensitivity (95% CI) | Specificity (95% CI) | PPV (95% CI) | NPV (95% CI) | F1-score |
| --- | --- | --- | --- | --- | --- | --- | --- |
| Confirmed BD | 26 | 25 | 88.5% [71.0%; 96.0%] | 97.3% [90.7%; 99.3%] | 92.0% [75.0%; 97.8%] | 96.0% [88.9%; 98.6%] | 0.902 |
| High probability | 12 | 10 | 25.0% [8.9%; 53.2%] | 92.0% [84.5%; 96.1%] | 30.0% [10.8%; 60.3%] | 90.0% [82.1%; 94.6%] | 0.273 |
| Some probability | 17 | 25 | 76.5% [52.7%; 90.4%] | 85.5% [76.4%; 91.5%] | 52.0% [33.5%; 70.0%] | 94.7% [87.1%; 97.9%] | 0.619 |
| No risk | 45 | 40 | 84.4% [71.2%; 92.3%] | 96.4% [87.7%; 99.0%] | 95.0% [83.5%; 98.6%] | 88.3% [77.8%; 94.2%] | 0.894 |

The highest category-specific performance was observed in the extreme classes, confirmed BD and no risk, whereas the two intermediate categories showed greater mutual confusion. This pattern supports the interpretation that the model is stronger at distinguishing clear presence or clear absence of bipolarity than at separating adjacent intermediate levels of risk.

## Notes

### Funding Statement

This study did not receive any specific grant from funding agencies in the public, commercial, or not-for-profit sectors. The artificial intelligence model evaluated in this study was developed by Arkangel AI within the context of an institutional collaboration with Clinica Montserrat.

### Author Declarations

The Institutional Research Ethics Committee of Clinica Montserrat - University Hospital (ICSN) gave ethical approval for this work (Act No. 228, December 5, 2025).

